# Association Between *Clostridioides difficile* Test Positivity and Incident Colorectal Cancer in a Multisite Hospital-Based Retrospective Cohort Analysis

**DOI:** 10.64898/2026.02.19.26346648

**Authors:** Samara Rifkin, Sean Anderson, Xingyu Chen, Kelly Gebo, Eili Klein, Nicholas O Markham, Matthew Robinson, Krishna Rao, Cynthia Sears

**Affiliations:** University of Michigan Medicine, Division of Gastroenterology/Hepatology, Ann Arbor, MI, USA; John D. Dingell VA Medical Center and Wayne State University School of Medicine, Division of Gastroenterology, Detroit, MI, USA; Johns Hopkins Medicine, Division of Infectious Diseases, Baltimore, MD, USA; Johns Hopkins University School of Medicine, Baltimore, MD, USA; Department of Emergency Medicine, Johns Hopkins University School of Medicine, Baltimore, MD, USA; Vanderbilt University Medical Center, Department of Medicine, Nashville, TN, USA; University of Michigan Medicine, Division of Infectious Diseases, Ann Arbor, MI, USA

**Keywords:** Colorectal cancer, *Clostridioides difficile*, microbiome

## Abstract

**Introduction:** Sporadic colorectal cancer (CRC) remains a significant driver of worldwide morbidity and mortality. Environmental factors associated with CRC are increasingly well-described and now include generalized colonic dysbiosis and individual enteric bacteria. *Clostridioides difficile* is one such species, with recent mouse model work suggesting prolonged exposure to *C. difficile* toxin B is conducive to colonic tumorigenesis. However, there is a dearth of real-world human evidence linking *C. difficile* infection and CRC.

**Methods:** Herein, we analyzed a multicenter, longitudinal, Electronic Health Record (EHR)-based dataset to test the association between *C. difficile* test positivity and the risk for incident CRC utilizing unadjusted and multivariable (controlled for clinical conditions independently associated with CRC development) Cox proportional hazard modeling to compare *C. difficile* exposed and non-exposed cohorts

**Results:** We found that individuals who tested recurrently positive for *C. difficile* had a significantly increased risk for incident CRC (aHR 2.05 [95% CI 1.27-3.29]) compared with those who tested positive only once (aHR 0.70 [0.45-1.10]) or never. Furthermore, we found potential trends that the effect of *C. difficile* test positivity on the risk for incident CRC was stronger amongst females compared with males.

**Importance:** These findings help translate emerging mouse model work on *C. difficile*-influenced colorectal tumorigenesis and lay groundwork for more substantial human investigations into this connection. These findings also may begin to help guide the personalized deployment of novel fecal microbiota-based therapies designed to interrupt the life cycle of *C. difficile* within the gut of human hosts and, potentially, prevent long-term health sequelae of chronic *C. difficile* infection.

## Introduction

Sporadic colorectal cancer (CRC) remains the second leading cause of cancer morbidity and mortality worldwide (1). Risk factors for sporadic CRC include both non-modifiable (e.g., older age, male sex, Black race) and modifiable (i.e., environmental) variables (2); the latter represents a critical area for study given the potential for disease prevention. Environmental factors contributing to CRC are complex, but there is emerging evidence that both generalized gut dysbiosis as well as individual enteric bacteria may contribute to colonic tumor formation (3). Dysbiosis and the overgrowth of certain potentially tumorigenic enteric bacteria are mediated by many factors including diet, tobacco/alcohol use, and obesity, all of which have been independently shown to be linked to CRC development (2). Importantly, oral antibiotic use, an established precipitant of gut dysbiosis, has been associated with an increased risk for CRC (4–6).

Oral antibiotics also strongly predispose patients to infection with *Clostridioides difficile,* a diarrheal pathogen transmitted via the fecal-oral route with spores that are commonly encountered in the environment, both in the community and healthcare settings. Asymptomatic carriage of *C. difficile* occurs in ∼10% of the “healthy” population (7) and is most frequent in infants and hospitalized patients (8). *C. difficile* infection (CDI) is defined as a clinically significant diarrheal illness that occurs upon germination of colonizing *C. difficile* spores and production of bacterial toxins. CDI occurs in over 500,000 people per year in the United States alone (9), and the clinical syndrome can vary from a mild diarrheal illness to fulminant and life-threatening colitis and sepsis. In recent years, female sex has been implicated as a risk factor for CDI (10), though the mechanism and long-term health consequences of this association remain unclear.

While CDI remains a significant public health concern given a rise in community-onset and recurrent cases in recent decades (11), little is known about the natural history of *C. difficile* colonization and persistence following initial diagnosis and treatment attempts. However, there is evidence that some patients can continue to shed spores for over a month after initial treatment of CDI, potentially leading to continued long-term exposure to spores after symptoms have abated (12). Unexpectedly, recent mouse model data showed that chronic colonization with toxigenic strains of *C. difficile* promotes colonic tumorigenesis (13). A notable nuance in this conclusion was the need for prolonged exposure to intraluminal toxin B (TcdB), the main virulence factor of *C. difficile* (14), for the pro-carcinogenic effect. The linkage between *C. difficile* infection and human CRC has not yet been thoroughly investigated. Taxonomic analyses of tissue in sporadic CRC have identified *C. difficile* enrichment (15) and several recent epidemiologic publications with limited study designs found conflicting results (16, 17). Specifically, these single-database studies relying on claims-based data found both increased and decreased CRC risk following an ICD-10 diagnosis of CDI, but these studies were likely limited by exposure misclassification and lack of temporal separation. In our study, we combined longitudinal data from two large institutions to test the association between a positive stool toxin assay for *C. difficile* and incident development of CRC among adults.

## 2.1 Methods

### 2.2 Study Design, Data Extraction, and Cohort Construction

We performed a retrospective cohort study utilizing longitudinal medical data from two large academic medical institutions. University of Michigan Medicine (MM) patient-level health data was obtained from a data repository containing demographics, anthropometric data, International Classification of Disease (ICD)-9 and ICD-10 diagnosis coding, laboratory testing values, and medication prescriptions. Johns Hopkins Medicine (JHM) patient-level health data was obtained from a repository collected under the Infectious Diseases Precision Medicine Center of Excellence, a dataset which contains ICD-10 diagnosis coding, microbiological test results, medication order and administration records, abstracted social and demographic history, and surgical and medical histories collected within EHR on every patient within JHM who ever received a positive microbiologic test result. Local IRB approval was obtained from each site to access data, which was fully de-identified prior to sharing between sites.

Each dataset was queried for all individuals 18 years of age and over who had completed stool-based testing for *C. difficile* within the timeframe captured by the respective database. For MM, the timeframe was between January 1, 2000 – August 2, 2023. For JHM the timeframe was between January 1, 2016 – August 15, 2024. We used the date of the first stool-based C. *difficile* testing to define the date of cohort entry (index date). For both sites, individuals were excluded if they were documented as deceased within 365 days after entry or if they did not have a follow-up encounter of any type within the system at least 365 days after their first *C. difficile* test. For JHH, ICD-10 codes were queried for the presence of any code compatible with a diagnosis of CRC (C18.0-7, C19, C20). For MM, prior CRC diagnoses were identified through the University of Michigan Rogel Cancer Center Registry. Individuals were excluded if their earliest date of any CRC diagnosis code was <365 days after entry *C. difficile* test (including all available information prior to cohort entry). We also excluded individuals with a history of total colectomy (44150, 44151, 44156–44158, 44210–4421) prior to baseline.

The individuals remaining in the dataset after application of these exclusion criteria were classified as *C. difficile* exposed if their first *C. difficile* test contained a positive result (including isolated NAT positivity as part of a two-step testing algorithm) and non-exposed if not. For individuals who initially tested negative and subsequently had a positive test for *C. difficile* prior to database exit, they were crossed over from unexposed to exposed groups; their follow-up time pre-positive test was counted as unexposed time, and their follow-up time post-positive test was counted as exposed time.

Finally, individuals who contributed any exposed follow-up time were categorized using a binary definition (never or ever) and nominal definition based on the number of positive *C. difficile* tests they had; if they had no positive *C. difficile* test they were categorized as CD=0; if only one positive test, CD=1; and if at least one subsequent *C. difficile* test had a positive result ≥30 days after a previous positive test, they were classified as CD >1. This was based on the histogram showing significant skew and zero inflation with most individuals having no exposure and most patients with *C. difficile* positivity having only one positive test, with very few having 2 or more positive tests. Similar to the negative to positive crossover as described above, in the nominal analysis, patients contribute time to multiple nominal exposure definitions with each additional infection (0, 1 or >1).

### 2.3 Covariates

Potential confounding risk factors were selected based on known clinical understanding of *C. difficile* and colon cancer pathogenesis. These variables included age at *C. difficile* testing, sex, obesity, diabetes mellitus (DM), preceding antibiotic use, preceding proton pump inhibitor (PPI) use, history of inflammatory bowel disease (IBD), and family history of colon cancer. Most of these metrics were assessed by querying the databases for ICD-10 codes compatible with these risk factors at the time of cohort entry. Age and sex were abstracted from demographic data for JHM and MM. At MM, BMI was abstracted from anthropometric data and obesity defined as BMI > 30 kg/m^2. At JHH, BMI data was not readily available, so ICD-10 codes for obesity were used instead. For both sites, preceding antibiotic use was determined by a query of medication records for all oral or intravenous antibiotics except for oral vancomycin or fidaxomicin, since these are treatments for CDI and not classic exposures, within 90 days prior to cohort entry. Dietary and behavioral factors (such as alcohol or tobacco use) are known risk factors for CRC, however given that these variables are not reliably coded into EHR data they were not included in this study’s analysis.

### 2.4 CRC Outcomes

The primary outcome was incident CRC >365 days after index date, which was ascertained using the MM cancer registry or by acquisition of a new ICD-10 code compatible with CRC (JHM). CRC diagnosis was also stratified into right (cecum, ascending colon and hepatic flexures), left (transverse colon, splenic flexure, descending colon and sigmoid), and rectal (rectum and rectosigmoid junction) location based on ICD-10 code if site-specific information was available (94% available for MM, 78% available for JHM).

### 2.5 Statistical Analysis

Individuals were compared by CD*-*exposure status (exposed vs. unexposed cohorts as above) using t-test and chi-square tests for continuous and categorical variables. We performed survival analysis to examine the effect of exposed versus unexposed status and incident CRC outcomes. Participants contributed time as either exposed or unexposed depending on whether the index lab test was positive or negative. Those who tested positive at any later date after an initial negative test began to contribute to exposed time on the date of the positive test. Participants were followed from the date of their entry *C. difficile* test until they were diagnosed with CRC, death, or censoring (last encounter date in the system), whichever occurred first.

We used unadjusted and multivariable multilevel random effects Cox proportional analysis to estimate the hazard ratio of incident CRC, right colon, left colon, and rectal cancer associated with exposed status (both binary exposure definition and nominal definition of exposed status) and 95% confidence intervals of colorectal cancer. The model incorporated random intercepts for site-level (MM and JHM) effects, allowing the relationship between exposed status and CRC risk to vary across sites. Predictors of incident CRC, estimates, and 95% confidence intervals (95% CI) were derived using a Cox model. The potential confounders included in the final model were selected based on prior epidemiologic literature on risk factors for CRC (2) and availability of variables at both clinical sites. The final multivariable Cox model included age, sex, race, DM, FHCRC, and personal history of IBD. We decided not to include antibiotics use into the model because a minority of individuals with C. difficile infection had prior antibiotic use and we assumed the variable must be missing information. Models were also stratified by sex. Interactions between CD status and sex were investigated by creating a cross product and applying a likelihood ratio test. Analyses were also conducted using 1-, 3-, and 5-year time-lag intervals between exposure and outcome, excluding CRC events occurring prior to each respective lag period using the MM cohort only. Proportional hazards assumptions were tested by including an interaction term between CD exposure and follow-up time in a Cox regression model. All analyses were conducted using R (version 4.4.2), with the survival package (version 3.8.3).

### 3.1 Results

### 3.2 Table 1: Cohort Characteristics & CRC Incidence

Overall, 100,181 individuals contributed 1,045,083 total person-years of follow-up. Seventy-nine percent of the individuals and ninety-one percent of the follow-up time were contributed by the MM cohort. At baseline, the median age of the MM cohort was 55 years, 9.5% were Black, and 54% were female whereas the JHM cohort was older with median age 62 years; 29% were Black; 57% were female. Patients from MM were more likely to have IBD, DM, and a FHCRC compared to JHM. Of all the patients tested for *C. difficile*, 15% and 24% tested positive in the MM cohort and the JHM cohort, respectively (17% overall positivity rate). Patients exposed to C. *difficile* were more likely to be female, older, carry a diagnosis of IBD or DM and have a recent history of antibiotic use. During the follow-up time, there were 254 incident CRC cases: 165 (65%) from MM and 89 (35%) from JHM; 41 (16%) of these incident CRC cases were recorded in the *C. difficile* exposed cohort. A breakdown of CRC by anatomic location is provided in Table 1. The overall incidence rate of CRC per 1,000 patient-years was 0.18 and 0.26 in the exposed and non-exposed cohorts, respectively.

**Table 1.** Baseline demographics of patients according to CD status, Michigan Medicine (2000–2023), Johns Hopkins (2016–2024)

|  | Total |  |  | Michigan Medicine |  |  | Johns Hopkins |  |  |
| --- | --- | --- | --- | --- | --- | --- | --- | --- | --- |
| Characteristic | No Hx of CD | Hx of CD | p-value<sup>2</sup> | No Hx of CD | Hx of CD | p-value<sup>2</sup> | No Hx of CD | Hx of CD | p-value<sup>2</sup> |
|  | N = 83,397<sup>1</sup> | N = 16,784<sup>1</sup> |  | N = 67,323<sup>1</sup> | N = 11,599<sup>1</sup> |  | N = 16,074<sup>1</sup> | N = 5,185<sup>1</sup> |  |
| Age | 56 (38, 69) | 57 (38, 70) | <0.001 | 54 (35, 67) | 55 (34, 67) | 0.1 | 62 (48, 73) | 62 (48, 74) | 0.6 |
| Sex |  |  | 0.01 |  |  | 0.6 |  |  | <0.001 |
| Female | 45,080 (54%) | 9,248 (55%) |  | 36,127 (54%) | 6,252 (54%) |  | 8,953 (56%) | 2,996 (58%) |  |
| Male | 38,313 (46%) | 7,528 (45%) |  | 31,192 (46%) | 5,339 (46%) |  | 7,121 (44%) | 2,189 (42%) |  |
| DM | 17,492 (21%) | 3,885 (23%) | <0.001 | 14,500 (22%) | 2,976 (26%) | <0.001 | 2,992 (19%) | 912 (18%) | 0.1 |
| IBD | 13,240 (16%) | 3,280 (20%) | <0.001 | 12,626 (19%) | 2,976 (26%) | <0.001 | 614 (3.8%) | 304 (5.9%) | <0.001 |
| FHCRC | 3,144 (3.8%) | 640 (3.7%) | 0.8 | 2,778 (4.1%) | 521 (4.5%) | 0.07 | 366 (2.3%) | 119 (2.3%) | >0.9 |
| Obesity |  |  | <0.001 |  |  | <0.001 |  |  | 0.2 |
| No | 47,229 (56%) | 11,624 (67%) |  | 31,416 (47%) | 6,221 (54%) |  |  |  |  |
| Yes | 17,571 (21%) | 3,334 (20%) |  | 16,803 (25%) | 3,110 (27%) |  | 768 (4.8%) | 224 (4.3%) |  |
| Unknown | 19,171 (23%) | 2,275 (13%) |  | 19,171 (28%) | 2,275 (20%) |  |  |  |  |
| Antibiotics | 36,240 (43%) | 8,965 (53%) | <0.001 | 25,879 (38%) | 5,690 (49%) | <0.001 | 10,361 (64%) | 3,275 (63%) | 0.1 |
| PPI | 19,746 (24%) | 4,096 (24%) | 0.05 | 17,428 (26%) | 3,577 (31%) | <0.001 | 2,318 (14%) | 519 (10%) | <0.001 |
| Race |  |  | <0.001 |  |  | <0.001 |  |  | 0.01 |
| Caucasian | 65,638 (79%) | 12,784 (76%) |  | 55,853 (83%) | 9,575 (83%) |  | 9,978 (61%) | 3,209 (62%) |  |
| Black | 10,877 (13%) | 2,709 (16%) |  | 6,377 (9.5%) | 1,223 (11%) |  | 4,500(28%) | 1,488 (29%) |  |
| Other | 6,787 (8.1%) | 1,272 (7.6%) |  | 5,093 (7.6%) | 803 (6.9%) |  | 1,694 (11%) | 469 (9.1%) |  |
| CRC | 213 (0.3%) | 41 (0.3%) | >0.9 | 139 (0.2%) | 26 (0.2%) | 0.8 | 74 (0.5%) | 15 (0.3%) | 0.1 |
| R colon | 64 (<0.1%) | 15 (<0.1%) | 0.6 | 53 (<0.1%) | 8 (<0.1%) | 0.9 | 11 (<0.1%) | 7 (0.1%) | 0.2 |
| L colon | 59 (<0.1%) | 11 (<0.1%) | 0.8 | 45 (<0.1%) | 10 (<0.1%) | 0.6 | 14 (<0.1%) | 1 (<0.1%) | 0.1 |
| Rectum | 65 (<0.1%) | 17 (0.1%) | 0.3 | 34 (<0.1%) | 8 (<0.1%) | 0.6 | 31 (0.2%) | 9 (0.2%) | 0.9 |
| PY | 4.0 (1.2, 7.7) | 4.0 (1.6, 7.2) | 0.007 | 3.9 (0.8, 8.7) | 4.1 (1.0, 8.9) | >0.9 | 4.07 (2.4, 5.8) | 3.97 (2.3, 5.7) | 0.005 |

<sup>1</sup> Median (Q1, Q3); n (%); <sup>2</sup> Welch Two Sample t-test; Pearson's Chi-squared test

### 3.3 Table 2: Multivariate Modeling for Association of *C. difficile* Exposure to CRC Risk

In both the MM and JHM cohorts, *C. difficile* exposure was not significantly associated with higher risk of CRC (adjusted HR, 1.43 [95% CI, 0.94-2.18] and adjusted HR, 0.62 [95% CI, 0.36 - 1.08], respectively). Similarly, when combined, *C. difficile* exposure was not significantly associated with higher risk of CRC (adjusted HR, 1.01 [95% CI, 0.72 - 1.42]). Stratified by sex, *C. difficile* exposure was not significantly associated with increased risk of CRC among women overall (adjusted HR, 1.22 [95% CI, 0.79 - 1.87]), nor or in either the MM cohort (adjusted HR, 1.67 [95% CI, 0.95 - 2.94]) nor the JHM cohort (adjusted HR, 0.85 [95% CI, 0.45 - 1.60]). For males, *C. difficile* exposure was significantly associated with reduced likelihood for CRC in the JHM cohort (adjusted HR, 0.30 [95% CI, 0.09 - 0.98]) but was not significantly associated with increased risk in the MM male cohort (adjusted HR, 1.20 [95% CI, 0.63 - 2.27]). Combined, the cohorts demonstrated no significant association in males. The interaction term of the cross product between *C. difficile* exposure and sex was also not significant (p=0.10).

**Table 2.**
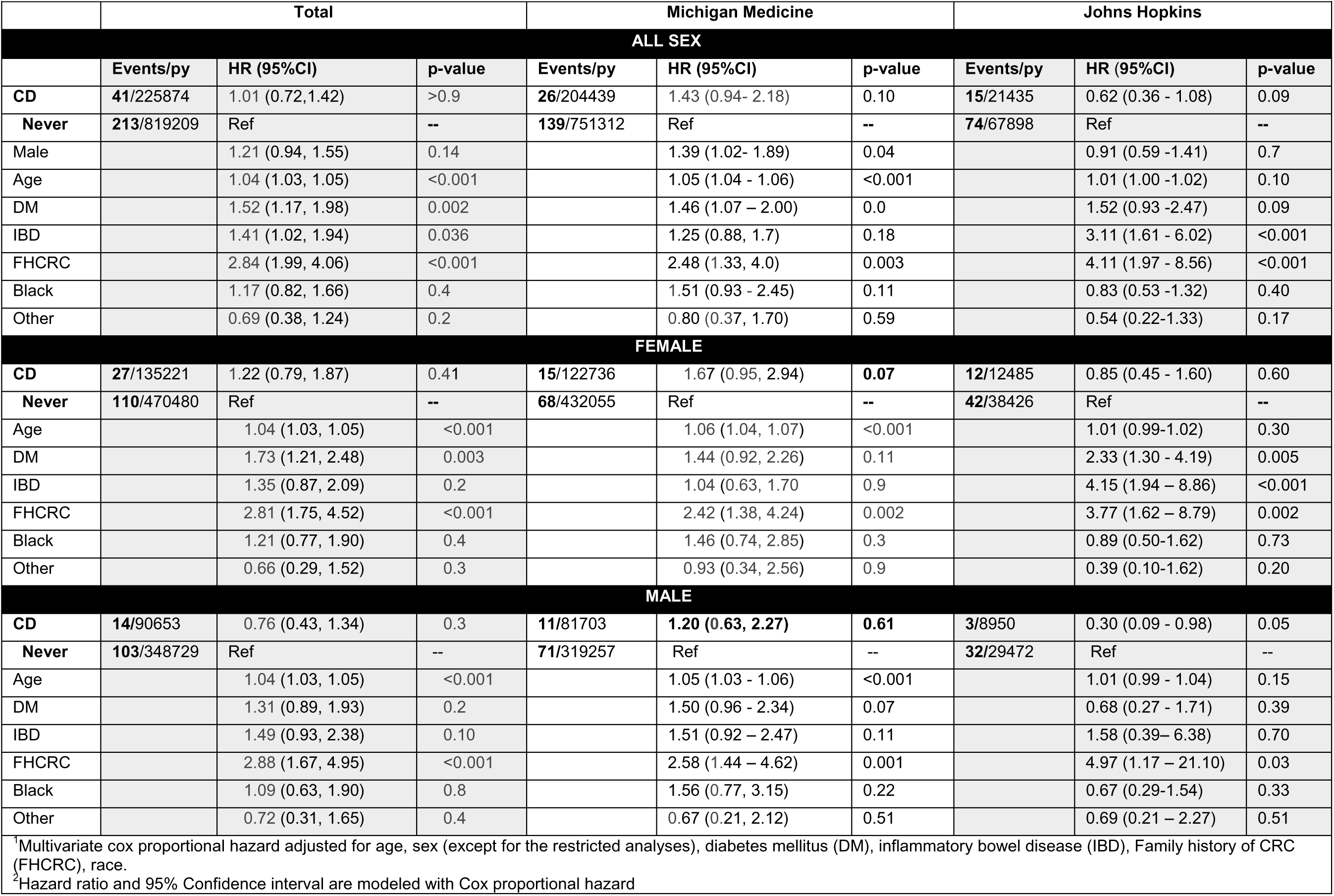
Association between any CD infection and colorectal incidence by sex, Michigan Medicine (2000–2023), Johns Hopkins (2016–2024)

|  | Total |  |  | Michigan Medicine |  |  | Johns Hopkins |  |  |
| --- | --- | --- | --- | --- | --- | --- | --- | --- | --- |
| ALL SEX |  |  |  |  |  |  |  |  |  |
|  | Events/py | HR (95%CI) | p-value | Events/py | HR (95%CI) | p-value | Events/py | HR (95%CI) | p-value |
| CD | 41/225874 | 1.01 (0.72,1.42) | >0.9 | 26/204439 | 1.43 (0.94- 2.18) | 0.10 | 15/21435 | 0.62 (0.36 - 1.08) | 0.09 |
| Never | 213/819209 | Ref | -- | 139/751312 | Ref | -- | 74/67898 | Ref | -- |
| Male |  | 1.21 (0.94, 1.55) | 0.14 |  | 1.39 (1.02- 1.89) | 0.04 |  | 0.91 (0.59 -1.41) | 0.7 |
| Age |  | 1.04 (1.03, 1.05) | <0.001 |  | 1.05 (1.04 - 1.06) | <0.001 |  | 1.01 (1.00 -1.02) | 0.10 |
| DM |  | 1.52 (1.17, 1.98) | 0.002 |  | 1.46 (1.07 – 2.00) | 0.0 |  | 1.52 (0.93 -2.47) | 0.09 |
| IBD |  | 1.41 (1.02, 1.94) | 0.036 |  | 1.25 (0.88, 1.7) | 0.18 |  | 3.11 (1.61 - 6.02) | <0.001 |
| FHCRC |  | 2.84 (1.99, 4.06) | <0.001 |  | 2.48 (1.33, 4.0) | 0.003 |  | 4.11 (1.97 - 8.56) | <0.001 |
| Black |  | 1.17 (0.82, 1.66) | 0.4 |  | 1.51 (0.93 - 2.45) | 0.11 |  | 0.83 (0.53 -1.32) | 0.40 |
| Other |  | 0.69 (0.38, 1.24) | 0.2 |  | 0.80 (0.37, 1.70) | 0.59 |  | 0.54 (0.22-1.33) | 0.17 |
| FEMALE |  |  |  |  |  |  |  |  |  |
| CD | 27/135221 | 1.22 (0.79, 1.87) | 0.41 | 15/122736 | 1.67 (0.95, 2.94) | 0.07 | 12/12485 | 0.85 (0.45 - 1.60) | 0.60 |
| Never | 110/470480 | Ref | -- | 68/432055 | Ref | -- | 42/38426 | Ref | -- |
| Age |  | 1.04 (1.03, 1.05) | <0.001 |  | 1.06 (1.04, 1.07) | <0.001 |  | 1.01 (0.99-1.02) | 0.30 |
| DM |  | 1.73 (1.21, 2.48) | 0.003 |  | 1.44 (0.92, 2.26) | 0.11 |  | 2.33 (1.30 - 4.19) | 0.005 |
| IBD |  | 1.35 (0.87, 2.09) | 0.2 |  | 1.04 (0.63, 1.70) | 0.9 |  | 4.15 (1.94 – 8.86) | <0.001 |
| FHCRC |  | 2.81 (1.75, 4.52) | <0.001 |  | 2.42 (1.38, 4.24) | 0.002 |  | 3.77 (1.62 – 8.79) | 0.002 |
| Black |  | 1.21 (0.77, 1.90) | 0.4 |  | 1.46 (0.74, 2.85) | 0.3 |  | 0.89 (0.50-1.62) | 0.73 |
| Other |  | 0.66 (0.29, 1.52) | 0.3 |  | 0.93 (0.34, 2.56) | 0.9 |  | 0.39 (0.10-1.62) | 0.20 |
| MALE |  |  |  |  |  |  |  |  |  |
| CD | 14/90653 | 0.76 (0.43, 1.34) | 0.3 | 11/81703 | 1.20 (0.63, 2.27) | 0.61 | 3/8950 | 0.30 (0.09 - 0.98) | 0.05 |
| Never | 103/348729 | Ref | -- | 71/319257 | Ref | -- | 32/29472 | Ref | -- |
| Age |  | 1.04 (1.03, 1.05) | <0.001 |  | 1.05 (1.03 - 1.06) | <0.001 |  | 1.01 (0.99 - 1.04) | 0.15 |
| DM |  | 1.31 (0.89, 1.93) | 0.2 |  | 1.50 (0.96 - 2.34) | 0.07 |  | 0.68 (0.27 - 1.71) | 0.39 |
| IBD |  | 1.49 (0.93, 2.38) | 0.10 |  | 1.51 (0.92 – 2.47) | 0.11 |  | 1.58 (0.39– 6.38) | 0.70 |
| FHCRC |  | 2.88 (1.67, 4.95) | <0.001 |  | 2.58 (1.44 – 4.62) | 0.001 |  | 4.97 (1.17 – 21.10) | 0.03 |
| Black |  | 1.09 (0.63, 1.90) | 0.8 |  | 1.56 (0.77, 3.15) | 0.22 |  | 0.67 (0.29-1.54) | 0.33 |
| Other |  | 0.72 (0.31, 1.65) | 0.4 |  | 0.67 (0.21, 2.12) | 0.51 |  | 0.69 (0.21 – 2.27) | 0.51 |

<sup>a</sup>Multivariate cox proportional hazard adjusted for age, sex (except for the restricted analyses), diabetes mellitus (DM), inflammatory bowel disease (IBD), Family history of CRC (FHCRC), race.
<sup>2</sup>Hazard ratio and 95% Confidence interval are modeled with Cox proportional hazard

### 3.4 Table 3: *C. difficile* Nominal Dose Effect & CRC Risk

We then assessed the association between multiple positive C. *difficile* tests and the risk of CRC. Of the 16,784 individuals in the exposed cohort, 3,815 (22%) were classified as multiply exposed (i.e., CDI > 1); 19 of the 41 CRC cases (46%) in the overall exposed cohort were identified in the CD >1 subgroup. Having two or more positive *C. difficile* assays spaced at least 30 days apart was significantly associated with an increased risk of CRC compared to non-exposed individuals (adjusted HR, 2.05 [95% CI, 1.27 - 3.29]) whereas a history of only one positive *C. difficile* assay did not show an increased risk (adjusted HR, 0.70 [95% CI, 0.45-1.10]). In the MM cohort, this positive association was stronger (adjusted HR, 2.97 [95% CI, 1.64 - 5.40]), while in the JHM cohort, the association was weaker and not statistically significant (adjusted HR, 1.23 [95% CI, 0.56–2.67]). When adjusting for the magnitude of *C. difficile* exposure, we found a significant association between exposure and CRC in females (adjusted HR for females, 2.43 [95% CI, 1.36 - 4.37]) but not in males (adjusted HR for males, 1.52 [95% CI, 0.66 - 3.49]). These associations were again stronger in the MM cohort.

**Table 3.** Association between nominal dose effect of CD exposure (0, 1 and > 1) and colorectal incidence by sex, Michigan Medicine (2000–2023), Johns Hopkins (2016–2024)

|  | Total |  |  | Michigan Medicine |  |  | Johns Hopkins |  |  |
| --- | --- | --- | --- | --- | --- | --- | --- | --- | --- |
| ALL SEX |  |  |  |  |  |  |  |  |  |
|  | Events/py | HR (95%CI) | p-value | Events/py | HR (95%CI) | p-value | Events/py | HR (95%CI) | p-value |
| CD=0 | 213/ 819209 | Ref | -- | 139/751311 | Ref | -- | 74/67898 | Ref | -- |
| CD =1 | 22/140775 | 0.70 (0.45, 1.10) | 0.12 | 14/ 124082 | 0.99 (0.57, 1.72) | >0.9 | 8/16693 | 0.44 (0.21, 0.90) | 0.03 |
| CD>1 | 19/85097 | 2.05 (1.27, 3.29) | 0.003 | 12/ 80357 | 2.97 (1.64, 5.40) | <0.001 | 7/4740 | 1.23 (0.56, 2.67) | 0.6 |
| Male |  | 1.21 (0.95, 1.55) | 0.13 |  | 1.39 (1.02, 1.90) | 0.035 |  | 0.92 (0.60, 1.41) | 0.7 |
| Age |  | 1.04 (1.03, 1.05) | <0.001 |  | 1.05 (1.04, 1.06) | <0.001 |  | 1.01 (1.00, 1.02) | 0.13 |
| DM |  | 1.50 (1.15, 1.96) | 0.002 |  | 1.45 (1.06, 1.99) | 0.021 |  | 1.50 (0.91, 2.45) | 0.11 |
| IBD |  | 1.38 (1.00, 1.91) | 0.047 |  | 1.22 (0.86, 1.73) | 0.3 |  | 3.10 (1.58, 6.09) | 0.001 |
| FHCRC |  | 2.83 (1.98, 4.04) | <0.001 |  | 2.47 (1.65, 3.70) | <0.001 |  | 4.04 (1.94, 8.41) | <0.001 |
| Black |  | 1.18 (0.83, 1.67) | 0.4 |  | 1.51 (0.93, 2.46) | 0.10 |  | 0.83 (0.51, 1.36) | 0.5 |
| Other |  | 0.70 (0.39, 1.25) | 0.2 |  | 0.80 (0.37, 1.71) | 0.6 |  | 0.54 (0.22, 1.35) | 0.2 |
| FEMALE |  |  |  |  |  |  |  |  |  |
| CD=0 | 110/470480 | Ref | -- | 68/432054 | Ref | -- | 42/38426 | Ref | -- |
| CD =1 | 14/81658 | 0.83 (0.48, 1.47) | 0.5 | 7/72076 | 1.02 (0.47, 2.22) | >0.9 | 7/ 9582 | 0.67 (0.30, 1.50) | 0.3 |
| CD>1 | 13/53563 | 2.43 (1.36, 4.37) | 0.003 | 8/50660 | 3.90 (1.85, 8.22) | <0.001 | 5/ 2902 | 1.35 (0.53, 3.42) | 0.5 |
| Age |  | 1.04 (1.03, 1.05) | <0.001 |  | 1.06 (1.04, 1.07) | <0.001 |  | 1.01 (0.99, 1.02) | 0.4 |
| DM |  | 1.70 (1.19, 2.44) | 0.004 |  | 1.43 (0.91, 2.25) | 0.12 |  | 2.29 (1.27, 4.15) | 0.006 |
| IBD |  | 1.32 (0.85, 2.04) | 0.2 |  | 1.00 (0.61, 1.64) | >0.9 |  | 4.12 (1.90, 8.94) | <0.001 |
| FHCRC |  | 2.79 (1.74, 4.48) | <0.001 |  | 2.41 (1.38, 4.23) | 0.002 |  | 3.69 (1.57, 8.66) | 0.003 |
| Black |  | 1.22 (0.78, 1.93) | 0.4 |  | 1.47 (0.75, 2.87) | 0.3 |  | 0.89 (0.49, 1.64) | 0.7 |
| Other |  | 0.67 (0.29, 1.53) | 0.3 |  | 0.94 (0.34, 2.59) | >0.9 |  | 0.40 (0.10, 1.65) | 0.2 |
| MALE |  |  |  |  |  |  |  |  |  |
| CD=0 | 103/348729 | Ref | -- | 71/ 319257 |  |  | 32/29472 | Ref | -- |
| CD =1 | 8/59117 | 0.55 (0.27, 1.14) | 0.11 | 7/ 52006 | 0.97 (0.45, 2.12) | >0.9 | 1/7111 | 0.12 (0.02 - 0.89) | 0.04 |
| CD>1 | 6/31535 | 1.52 (0.66, 3.49) | 0.3 | 4/ 29697 | 2.05 (0.74, 5.66) | 0.2 | 2/1838 | 0.92 (0.22, 3.86) | 0.99 |
| Age |  | 1.04 (1.03, 1.05) | <0.001 |  | 1.05 (1.03, 1.06) | <0.001 |  | 1.01 (1.00 - 1.04) | 0.05 |
| DM |  | 1.30 (0.88, 1.92) | 0.2 |  | 1.49 (0.96, 2.33) | 0.076 |  | 0.62 (0.25 - 1.57) | 0.30 |
| IBD |  | 1.47 (0.92, 2.36) | 0.11 |  | 1.49 (0.91, 2.44) | 0.12 |  | 1.30 (0.32 - 5.20) | 0.70 |
| FHCRC |  | 2.86 (1.67, 4.93) | <0.001 |  | 2.57 (1.44, 4.60) | 0.001 |  | 4.47 (1.07, 18.7) | 0.04 |
| Black |  | 1.09 (0.63, 1.91) | 0.8 |  | 1.56 (0.77, 3.15) | 0.2 |  | 0.63 (0.27, 1.46) | 0.30 |
| Other |  | 0.72 (0.32, 1.66) | 0.4 |  | 0.67 (0.21, 2.12) | 0.5 |  | 0.65 (0.20 - 2.17) | 0.50 |

<sup>1</sup>Multivariate cox proportional hazard adjusted for age, sex (except for the restricted analyses), diabetes mellitus (DM), inflammatory bowel disease (IBD), Family history of CRC (FHCRC), race.
<sup>2</sup>Hazard ratio and 95% Confidence interval are modeled with Cox proportional hazard

### 3.5 Table 4 and 5: Association Between Any *C. difficile* exposure and Nominal *C. difficile* Dose Effect and CRC Incidence by Anatomical Site

When evaluated by anatomic location broken down by proximal colon (i.e., right-sided), distal colon (i.e., left-sided), and rectal cancer, there was no significant association between binary *C. difficile* exposure and CRC. However, when the magnitude of *C. difficile* exposure was incorporated into the model, having multiple positive assays was significantly associated with proximal colon cancer (adjusted HR 3.28 [95% CI, 1.55 - 6.93]) and rectal cancer (adjusted HR 2.39 [95% CI, 1.14 - 5.03]) but not distal colon cancer (adjusted HR 2.26 [95% CI, 0.90 - 5.71]). For proximal colon cancer, the association was primarily driven by a strong positive correlation in the JHM cohort, while rectal cancer was more strongly associated with MM cohort.

**Table 4.**
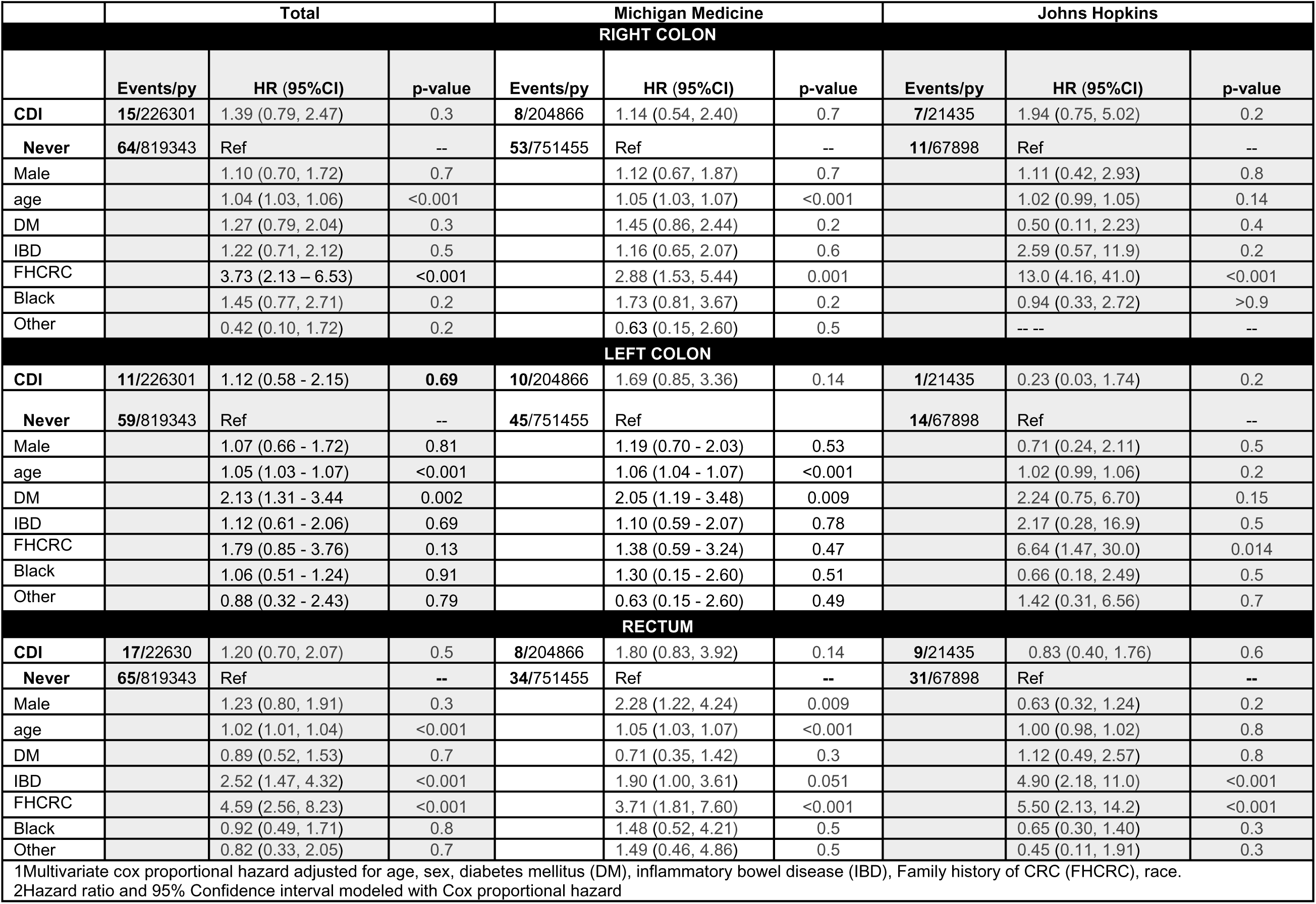
Association between any CDI exposure and colorectal incidence by site (R colon, L colon, Rectum), Michigan Medicine (2000–2023), Johns Hopkins (2016–2024)

**Table 5.** Association between nominal dose effect of CD infection (0, 1 and > 1) and colorectal incidence by anatomical site (R colon, L colon, Rectum), Michigan Medicine (2000–2023), Johns Hopkins (2016–2024)

|  | Total |  |  | Michigan Medicine |  |  | Johns Hopkins |  |  |
| --- | --- | --- | --- | --- | --- | --- | --- | --- | --- |
| RIGHT COLON |  |  |  |  |  |  |  |  |  |
|  | Events/py | HR (95%CI) | p-value | Events/py | HR (95%CI) | p-value | Events/py | HR (95%CI) | p-value |
| CD = 0 | 64/819343 | Ref | -- | 53/751445 | Ref | -- | 11/67898 | Ref | -- |
| CD =1 | 7/140915 | 0.84 (0.38, 1.85) | 0.7 | 4/124220 | 0.73 (0.27, 2.03) | 0.6 | 3/16695 | 1.09 (0.30, 3.93) | 0.9 |
| CD>1 | 8/85386 | 3.28 (1.55, 6.93) | 0.002 | 4/80645 | 2.55 (0.91, 7.10) | 0.074 | 4/4740 | 4.65 (1.46, 14.8) | 0.009 |
| Male |  | 1.11 (0.71, 1.73) | 0.7 |  | 1.13 (0.68, 1.88) | 0.6 |  | 1.12 (0.43, 2.95) | 0.8 |
| Age |  | 1.04 (1.03, 1.06) | <0.001 |  | 1.05 (1.03, 1.07) | <0.001 |  | 1.02 (0.99, 1.05) | 0.14 |
| DM |  | 1.25 (0.78, 2.01) | 0.4 |  | 1.44 (0.86, 2.42) | 0.2 |  | 0.47 (0.10, 2.08) | 0.3 |
| IBD |  | 1.18 (0.68, 2.06) | 0.5 |  | 1.13 (0.63, 2.02) | 0.7 |  | 2.45 (0.52, 11.5) | 0.3 |
| FHCRC |  | 3.71 (2.12, 6.51) | <0.001 |  | 2.88 (1.52, 5.44) | 0.001 |  | 12.5 (3.97, 39.2) | <0.001 |
| Black |  | 1.46 (0.78, 2.74) | 0.2 |  | 1.73 (0.81, 3.69) | 0.2 |  | 0.94 (0.32, 2.75) | >0.9 |
| Other |  | 0.42 (0.10, 1.73) | 0.2 |  | 0.63 (0.15, 2.61) | 0.5 |  | -- -- | -- |
| LEFT COLON |  |  |  |  |  |  |  |  |  |
| CD = 0 | 59/819343 | Ref | -- | 45/751445 | Ref | -- | 14/67898 | Ref | -- |
| CD =1 | 6/140915 | 0.79 (0.34, 1.84) | 0.6 | 5/124220 | 1.08 (0.43, 2.74) | 0.9 | 1/16695 | 0.30 (0.04, 2.32) | 0.3 |
| CD>1 | 5/85386 | 2.26 (0.90, 5.71) | 0.083 | 5/80645 | 3.89 (1.52, 9.95) | 0.004 | 0/4740 | -- -- | -- |
| Male |  | 1.07 (0.67, 1.73) | 0.8 |  | 1.20 (0.70, 2.04) | 0.5 |  | 0.71 (0.24, 2.10) | 0.5 |
| Age |  | 1.05 (1.03, 1.07) | <0.001 |  | 1.06 (1.04, 1.07) | <0.001 |  | 1.02 (0.99, 1.06) | 0.2 |
| DM |  | 2.11 (1.30, 3.41) | 0.002 |  | 2.03 (1.18, 3.47) | 0.010 |  | 2.26 (0.76, 6.76) | 0.14 |
| IBD |  | 1.11 (0.60, 2.04) | 0.7 |  | 1.07 (0.57, 2.01) | 0.8 |  | 2.17 (0.28, 17.0) | 0.5 |
| FHCRC |  | 1.78 (0.84, 3.75) | 0.13 |  | 1.38 (0.59, 3.23) | 0.5 |  | 6.72 (1.49, 30.3) | 0.013 |
| Black |  | 1.06 (0.51, 2.21) | 0.9 |  | 1.30 (0.55, 3.07) | 0.5 |  | 0.66 (0.17, 2.48) | 0.5 |
| Other |  | 0.88 (0.32, 2.45) | 0.8 |  | 0.64 (0.15, 2.62) | 0.5 |  | 1.42 (0.31, 6.53) | 0.7 |
| RECTUM |  |  |  |  |  |  |  |  |  |
| CD = 0 | 67/819343 | Ref | -- | 34/751445 | Ref | -- | 31/69145 | Ref | -- |
| CD =1 | 9/140915 | 0.83 (0.41, 1.69) | 0.6 | 5/124220 | 1.47 (0.57, 3.77) | 0.4 | 4/16901 | 0.73 (0.27, 2.03) | 0.6 |
| CD>1 | 8/85386 | 2.39 (1.14, 5.03) | 0.022 | 3/80645 | 2.93 (0.89, 9.69) | 0.78 | 5/4782 | 2.55 (0.91, 7.10) | 0.074 |
| Male |  | 1.24 (0.80, 1.93) | 0.3 |  | 2.29 (1.23, 4.25) | 0.009 |  | 1.13 (0.68, 1.88) | 0.6 |
| Age |  | 1.02 (1.01, 1.04) | <0.001 |  | 1.05 (1.03, 1.07) | <0.001 |  | 1.05 (1.03, 1.07) | <0.001 |
| DM |  | 0.88 (0.52, 1.50) | 0.6 |  | 0.70 (0.35, 1.41) | 0.3 |  | 1.44 (0.86, 2.42) | 0.2 |
| IBD |  | 2.47 (1.44, 4.25) | 0.001 |  | 1.86 (0.98, 3.55) | 0.059 |  | 1.13 (0.63, 2.02) | 0.7 |
| FHCRC |  | 4.55 (2.54, 8.14) | <0.001 |  | 3.70 (1.80, 7.57) | <0.001 |  | 2.88 (1.52, 5.44) | 0.001 |
| Black |  | 0.92 (0.49, 1.72) | 0.8 |  | 1.48 (0.52, 4.20) | 0.5 |  | 1.73 (0.81, 3.69) | 0.2 |
| Other |  | 0.82 (0.33, 2.06) | 0.7 |  | 1.49 (0.46, 4.85) | 0.5 |  | 0.63 (0.15, 2.61) | 0.5 |

1Multivariate cox proportional hazard adjusted for age, sex, diabetes mellitus (DM), inflammatory bowel disease (IBD), Family history of CRC (FHCRC), race.
2Hazard ratio and 95% Confidence interval modeled with Cox proportional hazard.

### 3.6 Table 6: Time-Lagged Associations Between Nominal *C. difficile* Exposure Dose Effect, and CRC Incidence

When evaluating the effect of time lag on the MM cohort, the overall association between *C. difficile* exposure on CRC incidence remained unchanged. An increased risk of CRC incidence among patients with more than one prior *C. difficile* test persisted at all lags times, although confidence intervals were wide. The association between a single prior positive C. difficile test and CRC incidence remained null.

**Table 6.** Time-Lagged Associations Between Nominal *C. difficile* Exposure Dose Effect and Colorectal Cancer Incidence, Michigan Medicine (2000–2023)

|  | 1-Year Lag |  |  | 3-Year Lag |  |  | 5-Year Lag |  |  |
| --- | --- | --- | --- | --- | --- | --- | --- | --- | --- |
| Characteristic | Events/py | HR (95% CI) | p-value | Events/py | HR (95% CI) | p-value | Events/py | HR (95% CI) | p-value |
| CD = 0 | 139/751312 | — | — | 96/70876 | — | — | 59/646257 | — | — |
| CD =1 | 14/124082 | 0.98 (0.57–1.70) | >0.9 | 10/116629 | 1.03 (0.53–1.97) | >0.9 | 8/05489 | 1.39 (0.66, 2.91) | 0.4 |
| CD>1 | 12/80358 | 2.58 (1.42–4.66) | 0.002 | 9/77852 | 2.70 (1.36–5.37) | 0.005 | 7/72765 | 3.62 (1.64, 7.99) | 0.001 |
| Male |  | 1.36 (1.00–1.86) | 0.049 |  | 1.41 (0.98–2.05) | 0.065 |  | 1.67 (1.05, 2.65) | 0.03 |
| Age |  |  |  |  |  |  |  | 1.05 (1.03, 1.06) | <0.001 |
| DM |  | 1.49 (1.09–2.05) | 0.014 |  | 1.54 (1.05–2.25) | 0.026 |  | 1.09 (0.67, 1.78) | 0.7 |
| IBD |  | 1.12 (0.79–1.59) | 0.5 |  | 0.92 (0.60–1.42) | 0.7 |  | 0.79 (0.45, 1.37) | 0.4 |
| FHCRC |  | 2.47 (1.65–3.70) | <0.001 |  | 2.82 (1.78–4.48) | <0.001 |  | 3.11 (1.80, 5.38) | <0.001 |
| Black |  | 1.44 (0.89–2.34) | 0.14 |  | 1.37 (0.77–2.46) | 0.3 |  | 1.03 (0.44, 2.40) | >0.9 |
| Other |  | 0.78 (0.37–1.67) | 0.5 |  | 0.62 (0.23–1.68) | 0.3 |  | 0.70 (0.22, 2.24) | 0.6 |

1Multivariate cox proportional hazard adjusted for age, sex, diabetes mellitus (DM), inflammatory bowel disease (IBD), Family history of CRC (FHCRC), race.
2Hazard ratio and 95% Confidence interval modeled with Cox proportional hazard.

## 4.1 Discussion

In this multicenter, retrospective cohort study, using EHR data, we identified an intriguing association between *C. difficile* assay positivity and the future risk for CRC. These results help advance the translation of mouse-model studies associating this ubiquitous pathogen with the pathophysiology of CRC. Most notably, when controlling for other variables established as independent predictors of CRC, we found that persistent or recurrent *C. difficile* exposure (defined as at least a second positive test at least 30 days following the first positive test) was significantly associated with the later development of CRC compared with those who only experienced a single positive test and those who never tested positive for *C. difficile* with at least 365 days of follow-up. This effect was consistent in both databases examined, although only statistically significant in the MM dataset. Additionally, we found trends suggesting a sex-based effect on *C. difficile-*associated CRC in that female sex seemed to strengthen the risk that *C. difficile* positivity had on eventual diagnosis of CRC.

These findings differ from other recent publications investigating similar questions. One study (11) utilizing data from the Florida Medicaid system found that a diagnosis of CDI was associated with an increased risk for CRC development in the subsequent 4 years. However, this study constructed their *C. difficile* cohort using ICD-based diagnosis claims for CDI as opposed to the use of *C. difficile* test results which we implemented in this present study. As a result, only 0.3% of the individuals analyzed were classified as exposed (compared with 17% in our study) which is likely a vast underestimation of true *C. difficile* exposure. Another more comprehensive study (12) used a national claims-based database to match CDI and non-CDI patients 1:1, based on ICD10 coding and National Drug Codes for antibiotics used to treat CDI. This study, interestingly, found opposite results, that CDI exposure was protective against CRC (except for obese patients, in whom CDI exposure increased the risk for CRC compared to non-exposed obese patients). That study, however, did not apply any time-based cutoffs for CRC diagnosis following CDI exposure, meaning that CRC diagnoses could have been essentially concurrent with CDI episodes; this calls any potential associations of *C. difficile* with the natural history of CRC into question. Furthermore, the matching was performed on many variables including age, sex, Charleson Comorbidity Index, CDI treatment, and obesity that may have inadvertently been overlooked as modifiers of the effect of *C. difficile* exposure on CRC. For example, we identified sex as a potential effect modifier, which would not have been accounted for due to the matching system utilized in that study.

Our detected relationship between persistent *C. difficile* exposure and CRC diagnosis is important for several reasons. First, these findings confirm similar results observed in the mouse model work previously mentioned (13); in these models, colorectal tumorigenesis was observed only in mice with prolonged *C. difficile* toxin production, and it disappeared in mice treated with vancomycin to eradicate the pathogen. Additionally, there is growing awareness of difficulty with *C. difficile* eradication despite appropriate antibacterial therapy and improvement in clinical symptoms in humans. In fact, a majority of recurrent CDI (rCDI) episodes are suspected to be caused by reactivation of non-eradicated bacterial spores as opposed to incident re-infection (14), and such chronic colonization and low-level toxin exposure could modify CRC risk. The incidence and consequences of failed eradication outside of a higher risk for rCDI are poorly studied in human hosts, but mouse models (13, 15) suggest that chronic enteric inflammation, dysbiosis, and possibly colorectal oncogenesis would be expected in susceptible hosts, potentially even in the absence of overt clinical symptoms of disease recurrence (i.e., subclinical infection). It is also known in human subjects that toxin production by *C. difficile*, sometimes at high levels, can occur in asymptomatic individuals (16).

Taken together, the above studies in mice and humans lead us to hypothesize that persistent colonization with toxigenic *C. difficile* leads to chronic, low-level toxin production and subclinical colon inflammation that contributes to CRC development in susceptible hosts. Persistent of *C. difficile* in human hosts following treatment attempts is a phenomenon suspected to be increasing in incidence since the early 2000’s due to dynamic changes in the predominating circulating strain of the pathogen to one with relatively high sporulation and decreased maximal toxin production potential (17), Additionally, our findings raise the possibility that host sex may influence the relationship between *C. difficile* exposure and CRC development. In our data, there appeared to be an association between *C. difficile* exposure of any magnitude and CRC development in females but not with males, however this did not reach statistical significance. A growing body of literature implicates female sex as a risk factor for both primary (10) and recurrent (18) CDI. Mechanisms purporting to explain this disparity remain unsolved but are hypothesized to involve a higher degree of healthcare (and specifically antibiotic prescription) engagement amongst females (19) and/or an influence of circulating estrogen on the modulation of *C. difficile* activity and gut microbiome composition (20).

The association between nominal *C. difficile* exposure and proximal colon or rectal (but not distal colon) cancer is suggested by our data, although statistically significant differences were not observed. The relatively small numbers of precisely coded CRC location diagnoses in our datasets make it difficult to hypothesize further about the potential anatomical influence of *C. difficile*-induced carcinogenesis. Interestingly, in the groundbreaking publication by Drewes et al (2022), mice who suffered from *C. difficile*-induced colon carcinogenesis seemed to exclusively develop distal colon tumors.

Such findings gain an added layer of relevance in an era of rapidly expanding options for the prevention of rCDI. Since 2022 two novel, live stool-based products have been approved by the FDA for rCDI prevention as quality-controlled, standardized alternatives for fecal microbiota transplantation. These products have been shown to significantly reduce rCDI rates in high-risk populations compared with those who receive antibiotics alone (21), although rates of *C. difficile* “eradication” following these therapies have not yet been extensively investigated and the products remain expensive (∼$9000-$17000/single administration). By advancing our understanding of who is at significant risk for long-term health complications (e.g., CRC) from persistent *C. difficile* toxin exposure through population-level studies such as this, we strive to personalize the decisions to deploy more aggressive interventions to eradicate *C. difficile* while balancing the cost of these interventions. Importantly, there is strong precedent for the identification, treatment, and confirmation of eradication of another chronic enteric pathogen, *Helicobacter pylori*, for the prevention of gastrointestinal cancers (22). While we acknowledge substantial differences in pathophysiology and the scientific evidence base between *H. pylori-* and *C. difficile-*induced oncogenesis, our findings raise the possibility that a similar conceptual framework may be applied to address the burden of CRC.

An important limitation of our study is the heterogeneity of follow-up time between the two databases integrated. In particular, the JHM database contains data from 2016-2024, a span of 8 years (compared to 23 years in the MM database). Conclusions regarding impacts of certain exposures on the natural history of CRC, which can take up to 10 years to develop (23), may be dampened. However, it is notable that median follow-up time is comparable between the two sites and that similar patterns of exposure effects on CRC are noted. Although the median follow-up in our study was relatively short (approximately 4-5 years) and shorter than the typical latency period for colorectal cancer development, we still identified a statistically significant association between *C. difficile* infection and CRC after applying a 1-year lag.

Sensitivity analyses using 3-year and 5-year lags produced similar results, although with wider confidence intervals. These findings suggest that the relationship may not be entirely dependent on long latency and could reflect effects on both early and later stages of the colorectal carcinogenesis pathway, including adenoma formation and progression. Inclusion in the JHM cohort required a positive microbiological test result at some point for entry, raising concern for underestimation of the non-exposed cohort. Additionally, the retrospective and EHR-based design of our study invites limitations inherent in this type of research. For example, although we required at least one recorded encounter for each individual at least one year after their entry into the cohorts, it is difficult to control for the quality of the follow-up encounter (e.g., physician visit versus imaging study) to ensure equitable risk for CRC diagnosis. We did not evaluate the effect of smoking and other behavioral risk factors including diet and physical activity, nor did we evaluate recent hospitalizations or surgeries. Furthermore, larger datasets are needed to further explore racial and anatomic subgroup differences.

Our study has several strengths compared to prior studies. Most importantly, our analysis method used *C. difficile* test results to construct cohorts as opposed to ICD10 coding for CDI, the latter of which likely underestimates the type of *C. difficile* exposure that mouse models suggest is biologically relevant for colorectal tumorigenesis (i.e., subclinical carriage). Additionally, our ability to stratify the nominal dose effect (i.e., multiply positive versus singly or never positive) of toxigenic *C. difficile* exposure by tracking persistently or recurrently positive test results without needing to rely on ICD coding for rCDI permits a high-resolution investigation into the effects of chronic exposure on CRC development, a novel technique for this question. Furthermore, the use of time-varying exposure models with cumulative dose updates over time enabled us to fully leverage the available data and improve estimate precision. To our knowledge, no prior studies have examined the CDI-CRC relationship using this type of survival analysis with time-varying exposure, which we consider a key strength of our study.

## 5.1 Conclusion

In summary, we have shown through this multicenter, retrospective cohort study that persistent exposure to toxigenic *C. difficile* is associated with the later development of CRC in U.S. adults, an effect that may be amplified by female sex. These findings advance mouse-model evidence of the role of *C. difficile* in effecting colorectal tumorigenesis and lay the groundwork for an enhanced awareness of and urgency in detecting and addressing chronic, subclinical *C. difficile* toxin production in individuals at heightened risk for CRC.

## Acknowledgements

**Financial Support:** S.M.A. is supported by the National Institutes of Health T32 A1007291. The content is solely the responsibility of the authors and does not necessarily represent the official views of the National Institutes of Health. K.R. is supported in part from an investigator-initiated grant from Merck & Co, Inc.; he has consulted for Seres Therapeutics, Inc., Rebiotix, Inc., Vedanta Biosciences, Inc., and Summit Therapeutics, Inc. N.M. is supported by VA CDA2 grant BX005699. The work is supported by the Bloomberg-Kimmel Institute for Immunotherapy, Cancer Research UK Cancer Grand Challenges Initiative OPTIMISTICC team grant C10674/A27140, and National Institutes of Health grant R01CA196845 (CLS), Page Foundation (SR), and a University of Michigan Pepper Center pilot grant AG024824 (SR). K.G. receives royalties from UpToDate, has served on a scientific advisory board for Shionogi and Pfizer (non-compensated), and has received personal consulting fees from Spark HealthCare, Premier HealthCare, Harrison Consulting and MedEd Learning.

## Abbreviations

CRC: Colorectal Cancer
EO-CRC: Early-Onset Colorectal Cancer
CDI: *Clostridioides difficile* Infection
rCDI: Recurrent *Clostridioides difficile* Infection
TcdB: *C. difficile* Toxin B
MM: University of Michigan Medicine
JHM: Johns Hopkins Medicine
ICD: International Classification of Disease
EMR: Electronic Medical Record
CD: *C. difficile*
IBD: Inflammatory Bowel Disease 95%
CI: 95% Confidence Interval
HR: Hazard Ratio
FHCRC: Family History of CRC
DM: Diabetes mellitus
PPI: Proton Pump Inhibitor

---

**Supplemental table 1.** Association between nominal dose effect of CD infection (0, 1 and > 1) and colorectal incidence by race, Michigan Medicine (2000–2023), Johns Hopkins (2016–2024)

| Characteristic | Total (Combined) |  |  | Michigan Medicine (MM) |  |  | Johns Hopkins (JHH) |  |  |
| --- | --- | --- | --- | --- | --- | --- | --- | --- | --- |
| CAUCASIAN |  |  |  |  |  |  |  |  |  |
|  | Events/py | HR (95%CI) | p-value | Events/py | HR (95%CI) | p-value | Events/py | HR (95%CI) | p-value |
| CD = 0 | 169/672839 | Ref | -- | 119/631607 | Ref | -- | 50/41232 | Ref | -- |
| CD =1 | 18/114318 | 0.77 (0.47, 1.26) | 0.3 | 11/104094 | 0.94 (0.51, 1.74) | 0.8 | 7/10224 | 0.55 (0.25, 1.21) | 0.13 |
| CD>1 | 13/69343 | 1.84 (1.04, 3.25) | 0.037 | 9/66232 | 2.66 (1.34, 5.28) | 0.005 | 4/3110 | 1.03 (0.37, 2.85) | >0.9 |
| Age |  | 1.04 (1.03, 1.05) | <0.001 |  | 1.05 (1.04, 1.06) | <0.001 |  | 1.01 (1.00, 1.03) | 0.089 |
| DM |  | 1.40 (1.04, 1.90) | 0.028 |  | 1.50 (1.06, 2.11) | 0.021 |  | 0.96 (0.47, 1.97) | >0.9 |
| IBD |  | 1.43 (1.01, 2.02) | 0.046 |  | 1.24 (0.85, 1.81) | 0.3 |  | 3.60 (1.66, 7.77) | 0.001 |
| FHCRC |  | 2.93 (1.99, 4.32) | <0.001 |  | 2.68 (1.75, 4.09) | <0.001 |  | 4.12 (1.64, 10.4) | 0.003 |
| Male |  | 1.29 (0.97, 1.70) | 0.076 |  | 1.40 (1.00, 1.96) | 0.05 |  | 1.02 (0.61, 1.70) | >0.9 |
| BLACK |  |  |  |  |  |  |  |  |  |
| CD = 0 | 35/10959 | Ref | -- | 15/75979 | Ref | -- | 20/19101 | Ref | -- |
| CD =1 | 6/10959 | 0.17 (0.02, 1.21) | 0.076 | 3/13053 | 0.55 (0.07, 4.17) | 0.6 | 0/4868 | -- | -- |
| CD>1 | 1/17921 | 3.50 (1.46, 8.37) | 0.005 | 1/9662 | 5.14 (1.43, 18.5) | 0.012 | 3/1297 | 1.83 (0.54, 6.23) | 0.3 |
| Age |  | 1.03 (1.01, 1.05) | 0.002 |  | 1.05 (1.02, 1.08) | <0.001 |  | 1.01 (0.98, 1.04) | 0.6 |
| DM |  | 1.72 (0.93, 3.18) | 0.084 |  | 1.26 (0.50, 3.17) | 0.6 |  | 2.44 (1.04, 5.72) | 0.04 |
| IBD |  | 1.15 (0.50, 2.65) | 0.7 |  | 1.23 (0.43, 3.50) | 0.7 |  | 2.61(0.59, 11.6) | 0.2 |
| FHCRC |  | 2.14 (0.76, 6.06) | 0.2 |  | 0.85 (0.11, 6.38) | 0.9 |  | 3.88 (1.12, 13.5) | 0.033 |
| Male |  | 0.93 (0.50, 1.74) | 0.8 |  | 1.43 (0.58, 3.56) | 0.4 |  | 0.66 (0.27, 1.60) | 0.4 |
| OTHER |  |  |  |  |  |  |  |  |  |
| CD = 0 | 9/51027 | Ref | -- | 5/43860 | Ref | -- | 4/7167 | — |  |
| CD =1 | 3/8603 | 2.38 (0.62, 9.06) | 0.2 | 2/7073 | 3.97 (0.76, 20.7) | 0.10 | 1/1530 | 1.13 (0.13, 10.2) | >0.9 |
| CD>1 | 0/5082 | -- | -- | 0/4751 | -- | -- | 0/331 | -- | -- |
| Age |  | 1.03 (1.00, 1.06) | 0.075 |  | 1.08 (1.02, 1.13) | 0.005 |  | 4.01 (0.63, 25.6) | 0.14 |
| DM |  | 1.89 (0.56, 6.31) | 0.3 |  | 0.99 (0.21, 4.55) | >0.9 |  | 0.00 (0.00, Inf) | >0.9 |
| IBD |  | 0.62 (0.08, 5.05) | 0.7 |  | 0.81 (0.10, 6.85) | 0.8 |  | 0.00 (0.00, Inf) | >0.9 |
| FHCRC |  | 2.56 (0.31, 20.8) | 0.4 |  | 2.61 (0.31, 22.0) | 0.4 |  |  |  |
| Male |  | 1.30 (0.42, 4.07) | 0.7 |  | 1.15 (0.25, 5.23) | 0.9 |  | 1.56 (0.26, 9.40) | 0.6 |

<sup>1</sup>Multivariate cox proportional hazard adjusted for age, sex, DM, IBD, FHCRC; <sup>2</sup>Hazard ratio and 95% Confidence interval modeled with Cox proportional hazard

## Author contributions

<u>Conceptualization</u>: S.R., S.M.A., N.O.M.,K.R., X.C., M.R., C.L.S.; <u>Methodology</u>: S.R, S.M.A., X.C., K.G., E.K., K.R., M.R., C.L.S. <u>Formal Analysis</u>: S.R., S.M.A., X.C. <u>Investigation</u>: S.R., S.M.A., X.C., M.R., K.R., C.L.S., <u>Resources</u>: S.R., K.G., E.K., K.R., M.R., C.L.S, <u>Writing – Original Draft:</u> S.R , S.M.A..; <u>Writing – Review &</u> <u>Editing</u>: all authors; <u>Visualization</u>: S.R., S.M.A., X.C., <u>Supervision</u>: C.L.S., M.R., K.R.; <u>Project</u> <u>Administration</u>: M.R., K.G., E.K., N.O.M., K.R., C.L.S.; <u>Funding Acquisition</u>: S.R., S.M.A., C.L.S., M.R.

## Data Availability

The data generated in this study are available within the article and its supplementary data files.

